# Continuous positive airway pressure face-mask ventilation to manage massive influx of patients requiring respiratory support during the SARS-CoV-2 outbreak

**DOI:** 10.1101/2020.06.01.20118018

**Authors:** S. Alviset, Q. Riller, J. Aboab, K. Dilworth, PA. Billy, Y. Lombardi, M. Azzi, L. Ferreira Vargas, L. Laine, M. Lermuzeaux, N. Memain, D. Silva, T. Tchoubou, D. Ushmorova, H. Dabbagh, S. Escoda, R. Lefrançois, A. Nardi, A. Ngima, V. Ioos

**Author notes:** **Corresponding author** Dr Vincent Ioos, MD, MSc, Mail: Service de Médecine Intensive Réanimation, Hôpital Delafontaine - 2, rue du, Docteur Delafontaine, 93200 SAINT DENIS - FRANCE.

## Abstract

**Background:** Since December 2019, a global outbreak of coronavirus disease (COVID-19) is responsible for massive influx of patients with acute respiratory failure in hospitals. We describe the characteristics, clinical course, and outcomes of COVID-19 patients treated with continuous positive airway pressure (CPAP) in a large public hospital in France.

**Method:** It is a single centre retrospective observational cohort. From 27th March to 23rd April, consecutive patients who had signs of respiratory failure or were unable to maintain an SpO_2_ > 90%, despite receiving 10 to 15 l/min of oxygen with a non-rebreather mask, were treated by CPAP with a face-mask unless the ICU physician judged that immediate intubation was indicated. The main outcomes under study were reasons for CPAP discontinuation and mortality.

**Results:** A total of 585 patients were admitted in Delafontaine hospital for severe COVID-19. ICU was quickly overwhelmed. Fifty-nine out of 159 (37%) patients requiring ICU care had to be referred to other hospitals. CPAP therapy was initiated in 49 patients and performed out of ICU in 41 (84%). SARS-CoV2 pneumonia was confirmed by PCR from respiratory tract in 39 (79%) patients and by thoracic CT scan in the remaining patients. CPAP was performed out of ICU in 41 (84%) cases. Median age was 65 years (IQR=54-71). Median duration of CPAP treatment was 3 days (IQR=1-5). Reasons for discontinuation of CPAP were intubation for invasive ventilation in 25 (51%) patients, improvement in 16 (33%), poor tolerance in 6 (12%) and death in 2 (4%). A decision not to intubate had been taken for the 2 patients who died while on CPAP.

**Conclusions:** Treatment with CPAP is feasible and safe in a non-ICU environment in the context of a massive influx of patients. One third of these patients with high oxygen requirements did not eventually need invasive ventilation.

**Key messages:** *What is the key question?:* What is the best respiratory support strategy to manage a massive influx of patients with hypoxemic respiratory failure despite high-flow oxygen delivered with a non-rebreather mask?

*What is the bottom line?:* Continuous positive airway pressure face mask ventilation delivered in non-ICU wards to patients who do not require immediate intubation is feasible and safe.

*Why read on?:* Face mask ventilation with CPAP should be considered as an option of respiratory support in the context of the on-going COVID-19 pandemic and limited availability of ICU beds.

## Background

The outbreak of the novel coronavirus disease 2019 (COVID-19) began in Wuhan, China in December 2019. Since then, it has rapidly spread around the world. As of May 19th, 2020, the WHO reported a total of 4 731 458 COVID-19 cases globally, with 6.67% mortality. In a large UK cohort, death from COVID-19 was strongly associated with being male, older age, deprivation, uncontrolled diabetes and severe asthma ^1^.

The nature of the pulmonary lesions triggered by SARS-CoV-2 is still a matter of debate. Some histopathological studies suggest that diffuse alveolar damage is not the single pattern ^2,3^ Disorders of the pulmonary circulation (thrombosis, endothelial injury) and organizing pneumonia may also be present. Many intensivists have observed that the classical clinical features of ARDS after intubation such as low pulmonary compliance are not found in all patients. A classification of mechanically-ventilated patients according to the driving pressure level after intubation has been proposed (L and H phenotypes) ^4,5^

In terms of clinical management, initial recommendations suggested early intubation and ARDS-type ventilator settings ^6^. Although some studies suggest a role for non-invasive ventilation in mild ARDS ^7-10^, invasive mechanical ventilation remains the standard of care, especially for severe cases. During the Chinese and European COVID-19 outbreaks, a number of critical care teams proposed using high flow nasal cannula or non-invasive ventilation at least for initial management ^11-14^ Optimal respiratory support for COVID-19 patients presenting with acute hypoxemic respiratory failure, however, remains unknown.

The district of Seine Saint Denis has been the worst affected area during the 2020 SARS-CoV-2 outbreak in Parisian region^15^. It is densely populated and has a high-deprivation index. From mid-March until end-April 2020, the Delafontaine Hospital, a large public hospital in Saint Denis, experienced a massive influx of patients requiring mechanical ventilation for acute respiratory failure due to COVID-19. During this period, the hospital in-patient bed capacity for non-ICU COVID-19 patients expanded to 210 beds. A total of 585 patients with SARS-CoV-2 infection were hospitalised. Despite increasing the number of intensive care beds from 18 to 32, the ICU was quickly overwhelmed. Fifty-nine (37%) out of 159 patients requiring ICU care had to be referred to other hospitals (Figure 1).

**Figure 1:**
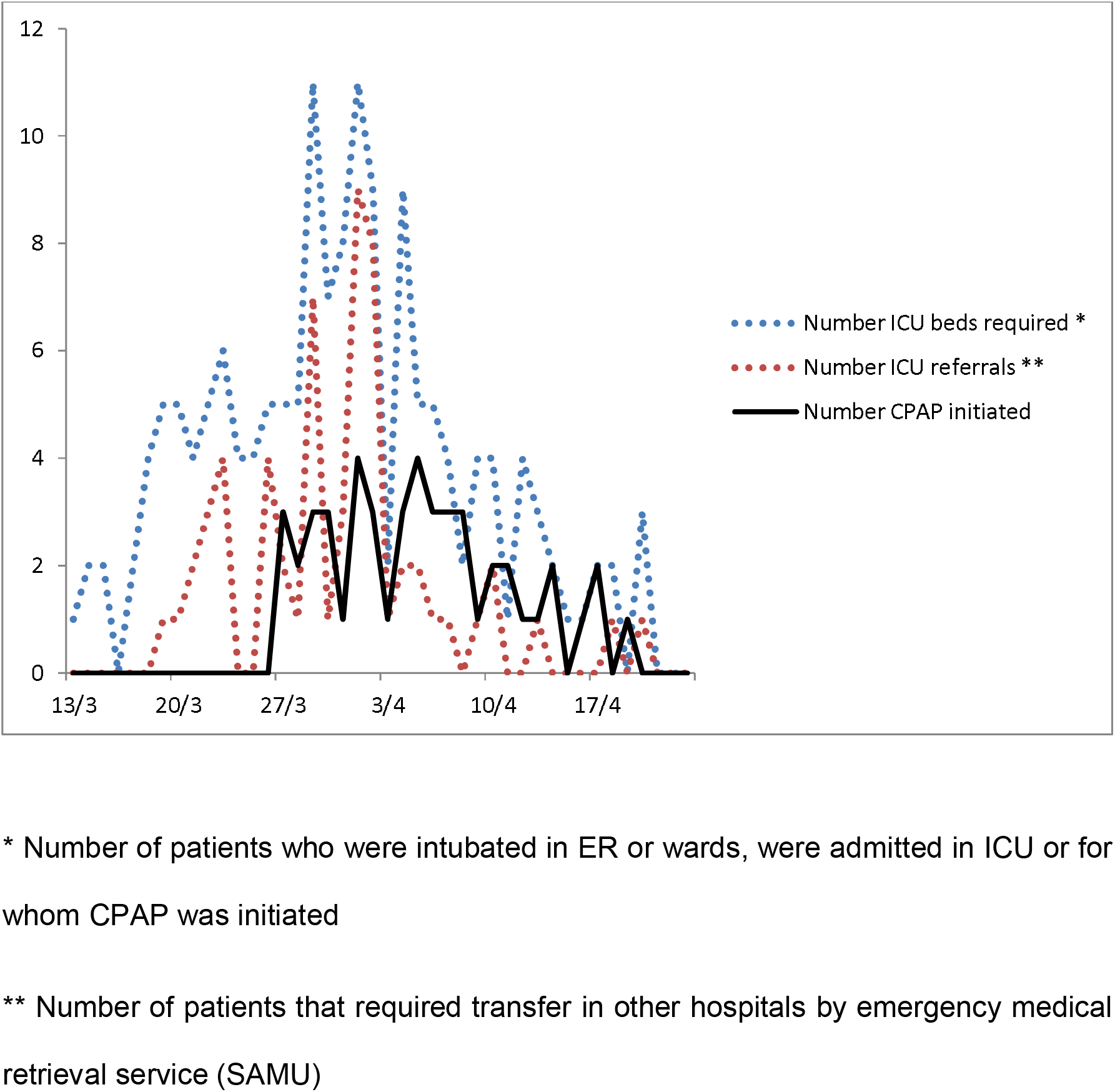
ICU patient load in Delafontaine Hospital during SARS-CoV-2 outbreak

To manage the flow of patients presenting from 27th march onwards, continuous positive airway pressure (CPAP) via face mask interface was considered in all patients with signs of respiratory failure despite 10 to 15 l/min of oxygen delivered by non-rebreather mask. In this single centre retrospective observational cohort study, we aim to describe the outcomes, in terms of clinical improvement without progression to intubation, need for intubation and mortality of patients supported with CPAP in our hospital during the SARS-CoV-2 outbreak.

## Materials and Methods

### Study design and participants

We reviewed the characteristics, clinical course and outcomes of all consecutive adults with proven COVID-19 treated with CPAP in ICU or in wards between 27th March and 23 April. During this 4 week-period, patients receiving 10-15 l/min oxygen through a non-rebreather mask who had clinical signs of respiratory failure or were unable to maintain an SpO2 > 90% were assessed for treatment with CPAP via facemask unless the ICU physician judged that immediate intubation was indicated. Every patient included in the study had a thoracic CT scan compatible with COVID-19 pneumonia and/or a positive SARS-CoV-2 PCR on naso-pharyngeal swab or broncho-alveolar lavage.

### Data collection

The following baseline patient characteristics were retrieved from patient electronic medical record: sex, age, comorbidities, body mass index (BMI), withholding / withdrawal of life-sustaining therapies, associated COVID-19 therapies (antivirals, steroids, immuno-modulating therapies, prone positioning), oxygen flow rate and SpO2 before and after starting CPAP treatment, duration of CPAP treatment, medical unit where CPAP treatment was performed, reasons for discontinuation of CPAP, duration of invasive mechanical ventilation, SAPS2 score for patients admitted in ICU, driving pressure and P/F ratio on first day of mechanical ventilation. The clinical outcomes (i.e. discharges from hospital, mortality) were recorded until the final day of follow-up on May 13^th^.

### CPAP therapy

CPAP of 5 to 10 cm H2O was delivered via a face mask dedicated to non-invasive ventilation (Performa Track®) with one of 2 types of CPAP valve (Boussignac™ or CPAP-O-two™) or alternatively, an ICU ventilator (Servo I® or Evita Infinity V500®). Treatment was undertaken in a medical ward, the emergency department (ED) short-stay unit or the ICU. An electrostatic heat and moisture exchanger filter (DAR™) was placed between the mask and the CPAP valve to prevent aerosolization of virus through expired gases. All patients were admitted to a single room with implementation of contact and airborne precautions; however some rooms were without a window. Medical and nursing staff in wards unfamiliar with non-invasive ventilation were trained by the intensivist who initiating the CPAP treatment. Patients received an initial prolonged session lasting at least 4 hours before being reassessed of their need of invasive mechanical ventilation. If the patient could be temporarily taken off CPAP without an immediate fall of SpO2 below 90% (on O2 15l/min via non-rebreather mask) or recurrence of clinical signs of acute respiratory failure, CPAP treatment was resumed for 2 hours every 4 hours. Progressive weaning of CPAP was performed according to clinical signs, pulse oximetry and arterial blood gases. If possible, patients were managed in the ICU (nurse/patient ratio 1:2). If no ICU bed was available (as in over 80% cases), patients with CPAP were shifted to the ED short-stay unit (8 beds) adjacent to the ICU (nurse/patient ratio 1:4) which allowed frequent re-evaluation of the patient’s state by the intensivist on call. In the eventuality of no bed availability in the ED short stay unit, CPAP treatment was instituted and managed in the medical ward were the patient had been admitted (nurse/patient ratio 1:7 during the outbreak). Ward patients on CPAP (and those with high O2 requirements) were systematically reviewed overnight by the duty resident responsible for the COVID-19 medical wards.

Non-invasive ventilation (NIV) with bi-level pressure modes was not used for three reasons: Firstly the number of ventilators available could not ensure surge capacity in the context of massive patient influx. Secondly, the increase in positive pressure during inspiration carries a greater risk of aerosolization of virus particles. The final reason was to keep pressure support ventilation as an option for pre-oxygenation before intubation in case it was indicated. Using bi-level pressure modes would have also required more intensive training of ward staff unfamiliar with NIV techniques.

### Statistics

No *a priori* statistical sample size calculation was performed. Sample size was equal to the number of patients treated during the study period. Quantitative values are expressed as the median (interquartile range, IQR), and qualitative values are presented as numbers (percentages). Univariate analysis was performed using Fisher exact test or Wilcoxon test, as appropriate. All tests were two-sided and a p value <0.05 was considered statistically significant. Because of alpha inflation due to multiple comparisons, findings should be interpreted as exploratory. A Cox hazard proportional model was fit for time to intubation, controlling for potential confounders in the cohort of 39 patients analysed. All variables available at baseline and associated with intubation in univariate analysis with a p-value <0.10 were selected. Variables selected are: CT-scan severity (<50% vs. ≥50 % of lung involved), SpO_2_ at the time of CPAP initiation, dose of anticoagulant (simple, double or curative) and time between hospital admission and CPAP use. All variables included in the Cox model were previously transformed to categorical variables. The cut-off value to construct these new variables were the median value for the 39 patients analysed. Missing data for SpO_2_ (n=3) at CPAP initiation were imputed based on the maximal bias assumption (i.e., low SpO_2_ in non-intubated patients and high SpO_2_ in intubated patients). Variables with more than 10 % missing values were not implemented in the multivariate analysis. The analyses were carried out using R version 3.6.2 (The R Project For Statistical Computing, Vienna, Austria; http://www.R-project.org).

### Ethics

The study was approved by the national ethics review board (CNRIPH - Commission Nationale des Recherches Impliquant la Personne Humaine) under the number 2020-A01396-33. The patients or their next-of-kin were informed by mail about the data collection process and their right to oppose. The database was declared to the Commission Nationale Informatique et Libertés (CNIL) under the number 2217928.

### No patient and public involvement

This research was done without patient involvement. Patients were not invited to comment on the study design and were not consulted to develop patient relevant outcomes or interpret the results. Patients were not invited to contribute to the writing or editing of this document for readability or accuracy.

## Results

Forty-nine consecutive patients were treated with CPAP between 27^th^ March and 23^rd^ April (Figure 2). Initiation of CPAP occurred throughout the entire study period and followed the epidemic curve (Figure 1). SARS-CoV-2 pneumonia was confirmed by PCR from upper or lower respiratory tract in 39 (79%) patients and by thoracic CT scan in the remaining patients. Twenty-six (53%) patients were eventually intubated and a total of 17 (34%) died.

**Figure 2:**
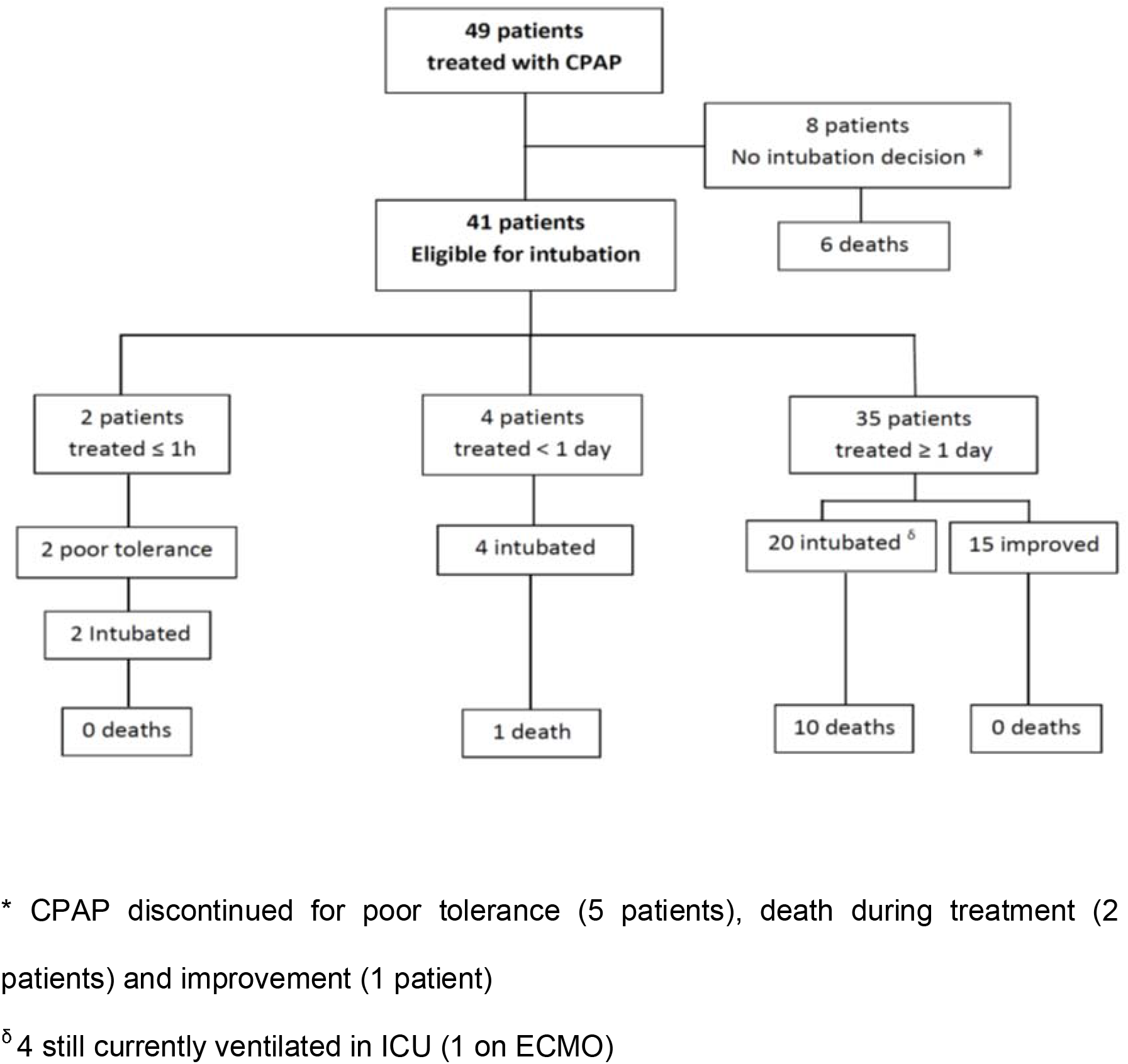
CPAP therapy - Patient flow diagram

Patients’ characteristics are presented in table 1. The median age was 65 years (IQR=54-71) and 36 (73%) were men. Forty-one (84%) patients had at least one comorbidity. The most frequent were hypertension (31 patients, 63%), obesity (13 patients, 34%) and diabetes (16 patients, 33%). The median duration of symptoms before hospital admission was 6 days (IQR = 5-9). Thoracic CT-scan at admission showed mild (10 to 25%), moderate (25 to 50%) or severe (>50%) lung involvement in 13 (27%), 23 (46%) and 13 (27%) patients respectively.

**Table 1.** Patients’ characteristics

| Characteristics | All patients (n=49) | Patients improved with CPAP (n= 15) * | Patients intubated after CPAP (n= 24) * | P value <sup>o</sup> |
| --- | --- | --- | --- | --- |
| Age in year <sup>ε</sup> | 65 (54-71) | 67 (53-68.5) | 62(54-69) | .79 |
| Age categories <sup>η</sup> |  |  |  |  |
| 0-39 yr | 0 | 0 | 0 |  |
| 40-49 yr | 5/49 (10%) | 1/15 (7%) | 3/24 (12.5%) | 1 |
| 50-59 yr | 11/49 (22%) | 4/15 (27%) | 6/24 (25%) | 1 |
| 60-69 yr | 19/49 (39%) | 8/15 (53%) | 9/24 (37.5%) | .51 |
| 70-79 yr | 10/49 (20%) | 2/15 (13%) | 5/24 (20%) | .69 |
| ≥80 yr | 4/49 (8%) | 0 | 1/24 (5%) | 1 |
| Female sex <sup>η</sup> | 13/49 (27%) | 4/15 (27%) | 5/24 (20%) | .71 |
| BMI distribution <sup>η</sup> |  |  |  |  |
| < 24,9 kg/m<sup>2</sup> | 10/38 (26%) | 3/11 (27%) | 6/21 (29%) | 1 |
| 25-29,9 kg/m<sup>2</sup> | 15/38 (40%) | 3/11 (27%) | 9/21 (43%) | .46 |
| 30-34,9 kg/m<sup>2</sup> | 8/38 (21%) | 3/11 (27%) | 4/21 (20%) | .67 |
| 35-39,9 kg/m<sup>2</sup> | 2/38 (5%) | 0 | 1/21 (4%) | 1 |
| ≥40 kg/m<sup>2</sup> | 3/38 (8%) | 2/11 (19%) | 1/21 (4%) | .27 |
| Comorbidities <sup>η</sup> |  |  |  |  |
| Any | 41/49 (84%) | 11/15 (73%) | 20/24 (83%) | .69 |
| Hypertension | 31/49 (63%) | 9/15 (60%) | 16/24 (67%) | .74 |
| Diabetes | 16/49 (33%) | 6/15 (40%) | 7/24 (29%) | .51 |
| Cerebrovascular disease | 3/49 (6%) | 1/15 (7%) | 2/24 (8%) | 1 |
| Coronary artery disease | 2/49 (4%) | 0 | 0 |  |
| Chronic renal failure | 5/49 (10%) | 2/15 (13%) | 3/24 (12.5%) | 1 |
| Chronic lung disease | 10 (20%) | 2/15 (13%) | 4/24 (17%) | 1 |
| Cancer | 1 (2%) | 0 | 0 |  |
| Immunodeficiency | 0 | 0 | 0 |  |
| Delay between symptoms and hospital admission <sup>ε</sup> | 6 (5-9) | 6 (5.5-9.5) | 6 (5-8.25) | .76 |
| SARS-CoV-2 PCR positivity <sup>η</sup> | 39/47 (83%) | 10/14 (71%) | 21/24 (88 %) | .39 |
| Thoracic CT at admission <sup>η</sup> |  |  |  |  |
| 10-25% | 13/49 (27%) | 7/15 (47%) | 4/24 (17%) | .07 |
| 25-50% | 23/49 (46%) | 6/15 (40%) | 13/24 (54%) | .75 |
| >50% | 13/49 (27%) | 2/15 (13%) | 7/24 (29%) | .15 |

\* Patients excluded for analysis (n=10): withdrawal/limitations of life-sustaining therapies (n=8), <1 hour CPAP treatment (n=2)
<sup>ε</sup> Median (IQR)
<sup>η</sup> Number / total number (%)

Modalities of CPAP therapy and associated interventions are described in table 2. CPAP was performed out of ICU in 41 (84%) cases. Median duration of CPAP therapy was 3 days (IQR=1-5). Reasons for discontinuation of CPAP were intubation for invasive ventilation in 25 (51%) patients, improvement in 16 (33%), poor tolerance in 6 (12%) and death in 2 (4%). A decision not to intubate had been taken with the patient and their family for the 2 patients who died while on CPAP. All patients received at least once daily prophylactic anticoagulation. Twice daily (thus double dose) prophylactic anticoagulation, typically enoxaparin 40mg every 12 hours, was administered in 19 (39%) patients while 14 (29%) received therapeutic anticoagulation. Hydroxychloroquine was administered in 17 (35%) patients, Lopinavir/Ritonavir in 4 (8%), corticosteroids in 28 (57%) and Anakinra in 7 (14%). Awake prone positioning was used in 7 (14%) patients. Two of those were eventually intubated.

**Table 2.** CPAP therapy and other interventions (before or during CPAP period)

|  | All patients | Patients improved with CPAP (n= 15)* | Patients intubated after CPAP (n= 24)* | P value |
| --- | --- | --- | --- | --- |
| Time between admission and CPAP initiation in days <sup>ε</sup> | 3 (1-5) | 4 (4-6.5) | 2 (1-5) | **.04** |
| Care zone of initiation of CPAP |  |  |  |  |
| ICU | 8/49 (16%) | 2/15 (13%) | 5/24 (21%) | .69 |
| ED short stay unit | 29/49 (59%) | 9/15 (61%) | 16/24 (67%) | .74 |
| COVID-19 ward | 12/49 (25%) | 4/15 (26%) | 3/24 (12%) | .69 |
| CPAP device |  |  |  |  |
| Boussignac™ valve | 41/49 (84%) | 11/15 (74%) | 20/24 (83%) | .69 |
| CPAP-O-two™ valve | 3/49 (6%) | 2/15 (13%) | 1/24 (4%) | .55 |
| ICU ventilator | 5/49 (10%) | 2/15 (13%) | 3/24 (13%) | 1 |
| Oxygen flow rate before CPAP initiation |  |  |  |  |
| 15L/min | 42/47 (89%) | 11/15 (73%) | 22/22 (100%) | .05 |
| 11-12L/min | 5/47 (11%) | 4/15 (27%) | 0 | .05 |
| SpO2 in % before CPAP initiation <sup>ε</sup> | 92 (90-95) n=44 | 95 (92.5-95.5) n=15 | 92 (90-93) n=21 | **.02** |
| Respiratory rate per min before CPAP <sup>ε</sup> | 36 (30-40) n=36 | 38 (29-40) n=13 | 32 (30-38) n=19 | .47 |
| Oxygen flow on CPAP in L/min | 25 (23-25) n=21 | 25 (23-26) n=5 | 25 (25-26) n=12 | 1 |
| SpO2 in % on CPAP <sup>ε</sup> | 97 (94-98) n=29 | 98 (96-98) n=9 | 96 (93-98) n=15 | .14 |
| Respiratory rate per min <sup>ε</sup> | 34 (29-37) n=23 | 29 (23-32) n=6 | 36 (30-37) n=14 | .46 |
| CPAP duration in days <sup>ε</sup> | 3 (1-5) | 4 (3-7) | 2 (2-3) | **.002** |
| <1h <sup>η</sup> | 4/49 (8%) | - | - |  |
| 1h - 1day <sup>η</sup> | 6/49 (12%) | 0 | 4/24 (17%) | .15 |
| 1-5 days <sup>η</sup> | 30/49 (61%) | 10/15 (67%) | 19/24 (79%) | .46 |
| >5 days <sup>η</sup> | 9/49 (18%) | 5/15 (33%) | 1/24 (4%) | **.02** |
| Other interventions |  |  |  |  |
| Antibiotics<sup>ο</sup> | 45/49 (92%) | 13/15 (87%) | 23/24 (96%) | .55 |
| Lopinavir/ritonavir | 4/49 (8%) | 1/15 (7%) | 1/24 (4%) | 1 |
| Corticosteroids | 28/49 (57%) | 9/15 (60%) | 11/24 (46%) | .30 |
| Anakinra | 7/49 (14%) | 2/15 (13%) | 3/24 (13%) | 1 |
| Hydroxychloroquine | 17/49 (35%) | 6/15 (40%) | 9/24 (38%) | 1 |
| Therapeutic anticoagulation | 14/49 (29%) | 3/15 (20%) | 7/24 (29%) | .71 |
| Twice daily (double dose) prophylactic anticoagulation | 19/49 (39%) | 9/15 (60%) | 6/24 (25%) | **.04** |
| Once daily (single dose) prophylactic anticoagulation | 16/49 (32%) | 3/15 (20%) | 11/24 (46%) | .17 |

\* Patients excluded for analysis (n=10): withdrawal/limitations of life-sustaining therapies (n=8), <1 hour CPAP treatment (n=2)
<sup>ε</sup> Median (IQR)
<sup>η</sup> Number / total number (%)
<sup>ο</sup> Antibiotics: Amoxicillin-clavulanic acid, Spiramycin, Azithromycin, Erythromycin, third generation cephalosporin, Piperacillin-tazobactam, Cefepim

Eight patients had a withdrawal/limitation of life-sustaining therapies (decision “do-not intubate”). Of the 41 other patients, 2 failed to tolerate CPAP resulting in its discontinuation within less than one hour. We did not consider these patients as being significantly treated, which left 39 study patients suitable for analysis of outcome. Fifteen (38%) patients out of 39 showed sustained clinical improvement with CPAP therapy and never required intubation. The other 24 (62%) patients eventually required invasive ventilation (Figure 2). For these 24 patients, median time from CPAP initiation to intubation was 1 day (IQR=1-2), median P/F immediately after intubation was 100 (IQR=80-139), and median duration of mechanical ventilation was 16 days (IQR=10-23).

Patients who improved with CPAP were compared to the group who ultimately progressed to needing intubation. Characteristics regarding age, sex, comorbidities and disease presentation were similar in both groups. Patient who improved on CPAP were treated later in their hospital stay, had higher oxygen saturation before CPAP initiation, longer duration of CPAP and received more often concomitant double dose prophylactic anticoagulation. A cox proportional hazard model was made to assess for confounding factors, variables associated with the risk of intubation in univariate analysis (p value < 0.10) were selected. Thoracic CT-scan severity, delay between hospital admission and use of CPAP or dosage of anticoagulant treatment was not associated with the risk of intubation. However, low oxygen saturation just before initiation of CPAP was associated with a higher risk of intubation (Figure 3).

**Figure 3.**
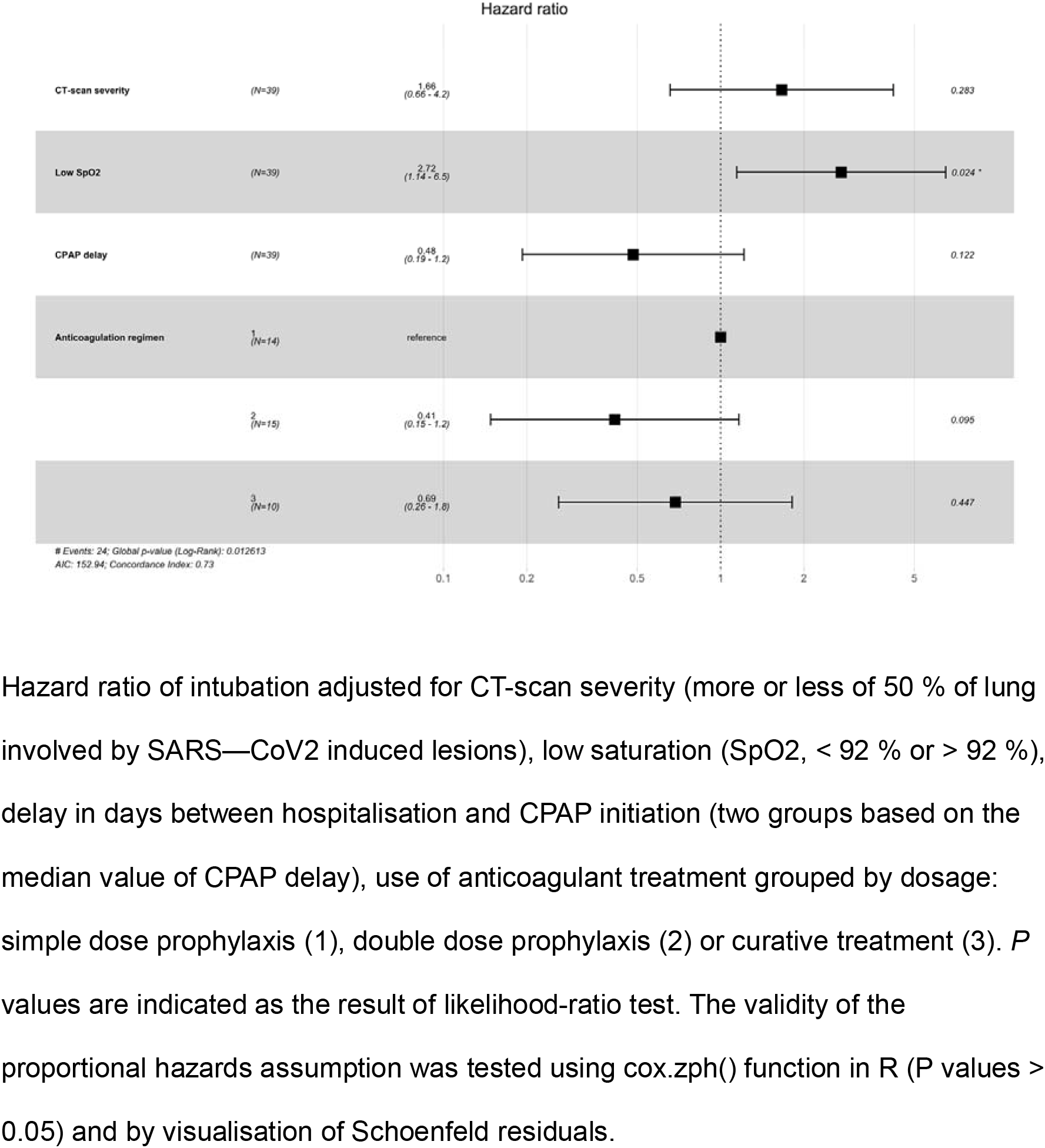
Factors associated with intubation Hazard ratio of intubation adjusted for CT-scan severity (more or less of 50 % of lung involved by SARS—CoV2 induced lesions), low saturation (SpO_2_, < 92 % or > 92 %), delay in days between hospitalisation and CPAP initiation (two groups based on the median value of CPAP delay), use of anticoagulant treatment grouped by dosage: simple dose prophylaxis (1), double dose prophylaxis (2) or curative treatment (3). *P* values are indicated as the result of likelihood-ratio test. The validity of the proportional hazards assumption was tested using cox.zph() function in R (P values > 0.05) and by visualisation of Schoenfeld residuals.

Eleven (42%) of the 26 intubated patients had a fatal outcome. Median SAPS 2 score of ventilated patients was 57 (IQR=38-64), resulting in a standardized mortality ratio of 0.88. At the time of final follow up, 17/49 (35%) patients had died, 15 (30%) were discharged (14 from the group of patients who improved with CPAP), 17 (35%) were still hospitalized (13 from the group of patients who were intubated after CPAP), of whom 4 (8%) remained on mechanical ventilation and 1 on ECMO.

## Discussion

This single center retrospective observational study describes the largest cohort to date of COVID-19 adult patients treated with CPAP via face mask.

The main purpose of using CPAP was to facilitate management of the patient flow of those potentially requiring invasive ventilation during this SARS-CoV-2 outbreak. CPAP via face mask does not require a ventilator, and could thus be instituted and run on non-ICU wards. This proved critical in this particular instance. Sixteen (33 %) patients improved with CPAP treatment, and eventually did not go on to require invasive ventilation though they were very hypoxemic (11 (73%) of them required 15L/min oxygen). Other than 2 patients with a do-not-intubate order, no death occurred during CPAP therapy. Mortality was 42.3% in the patient group requiring intubation. This was related to the severity of illness (median SAPS2 score of 57) but may be also due to difficulties in maintaining the quality of ICU care during a crisis situation. Mortality data for COVID-19 patients on invasive ventilation is still scarce, but early reports from China and the US showed mortality rate ranging between 76 and 97%^16–19^. In Bergamo (Italy), an overall mortality of 61.2% was reported among 99 patients necessitating ventilator support by Helmet CPAP (84%), non-invasive (8%) or invasive (8%) ventilation in the setting of the SARS-CoV-2 outbreak ^20^ In a larger study in Lombardy, of 1591 ICU patients admitted with COVID-19, 72% were mechanically ventilated while 9% were treated with non-invasive ventilation. Mortality rate was 26% at the end of the study period with 920 (58%) patients were still in ICU^21^. In Vancouver (Canada), the mortality rate among 117 patients (of which 62.3% were on mechanical ventilation) admitted in 6 ICUs was 15.3%, while 10.3% remained in the ICU ^22^.

Use of CPAP has already been reported in outbreaks of acute severe respiratory infection such as the 2003 SARS epidemic. However it was used in patients with less profound hypoxemia than in our cohort (5 to 6 l/m), and a lower percentage (10 to 30%) of patients required intubation ^23,24^

This study has several limitations. Firstly, due to its retrospective design, we were unable to collect additional data that could have contributed to a better understanding of the role of CPAP in managing hypoxemic respiratory failure in COVID-19. Data on actual pressure levels delivered to each patient and the number of hours per day of CPAP therapy could not be retrieved. In addition, it was not possible to ascertain in all patients whether vital signs (SpO2, respiratory rate) and arterial blood gases were taken while on CPAP or while on non-rebreather mask. Finally, the absence of a control group does not allow us to make any firm conclusion on the role of CPAP in avoiding intubation.

Secondly, due to small sample size, the observed effect of CPAP in avoiding invasive mechanical ventilation within a sub-group of patients could be biased by concomitant treatments (drugs and/or prone positioning during spontaneous breathing) administered to spontaneously breathing-patients. Another possible limiting factor are the higher oxygen flow rates used during CPAP therapy compared with nonrebreather masks (20 to 30 l/m versus 15 l/m), which could have contributed to the clinical improvement of patients, by increasing the FiO2 delivered. It is therefore difficult to conclude that patients improved uniquely because of CPAP.

Third and finally, contamination of health workers was not evaluated. Expired gases dispersion during CPAP seems to be limited if there is good mask interface fitting ^25^, but leaks do occur incidentally and non-invasive ventilation is considered an aerosol-generating procedure. Potential benefit from CPAP via face mask needs to be weighed against the risk of contamination for health care workers, especially in settings were infection prevention and control precautions are difficult to maintain. Choosing the appropriate interface is critical to decrease leaks and minimize aerosolization and there may be some advantages to select full face masks. Helmet is another option but is more difficult to handle in a non-ICU setting.

Patients with profound hypoxemia and high respiratory rate who are treated with CPAP may be exposed to self-induced lung injury. We attempted to collect the values for driving pressures immediately after intubation, but these data was unfortunately only available in a few cases. This should be investigated in further studies.

On one hand, CPAP treatment may have in fact triaged a group of patients who were less severe. This selection effect is suggested by the higher levels of SpO2 at initiation of CPAP in the group of patients who improved compared with the group of patients who progressed to intubation. CPAP therapy could have potentially worsened the condition of patients whose intubation was then effectively delayed. The high SAPS2 scores of the intubated patients in the study provide some evidence to this effect.

On the other hand, there may be some advantages of using CPAP even for patients who are subsequently intubated. CPAP prior to intubation may reduce the duration of mechanical ventilation, thereby decreasing the risk of ventilator acquired pneumonia and exposure to ventilator induced lung injury.

The role of face mask CPAP ventilation in managing acute hypoxemic respiratory failure in COVID-19 patients warrants further investigation in larger prospective studies. The simplicity and practicality of this technique in a number of contexts, including massive patient influx and resource limited settings, is appealing^26^. However, the likely increased risk of contamination of heath care workers, notably if personal protective equipment is inadequate, must be taken in account. CPAP could also be considered as a first-line respiratory support strategy in less hypoxemic patients without significant respiratory failure in association with other strategies to improve oxygenation, such as awake prone positioning^27,28^.

In conclusion, we found treatment with face mask CPAP to be feasible and safe in a non-ICU environment and in the context of a massive influx of patient. It was useful to post-pone intubation and to manage the flow of patient requiring invasive ventilation. We also found, that among patients who have low SpO_2_ and /or signs of respiratory failure while on 15l/min O2 via non rebreather mask about one third eventually did not need invasive ventilation.

## Data Availability

The data that support the findings of this study are available on request from the corresponding author, VI. The data are not publicly available due to their containing information that could compromise the privacy of research participants.

## Acknowledgements

Author Contributions: Ioos had full access to all the data in the study and takes responsibility for the integrity of the data and the accuracy of the data analysis.

## Concept and design

Ioos, Alviset

## Acquisition, analysis, or interpretation of data

Ioos, Alviset, Riller,

## Critical revision of the manuscript for important intellectual content

Dilworth, Billy, Azzi, Ferreira, Laine, Lermuzeaux, Memain, Silva, Tchoubou, Ushmorova, Dabbagh, Escoda, Lefrançois, Nardi, Ngima

## Statistical analysis

Riller, Lombardi

## No funding was required for this study

CPAP devices were purchased through the regular hospital supply.

## Supervision

Ioos

## Disclaimer

The findings and conclusions in this report are those of the authors and do not necessarily represent the official position of the Centers for Disease Control and Prevention.

## Additional Contributions

We thank the doctors, residents and nursing staff of the Delafontaine Hospital who managed patients under CPAP therapy during the SARS-CoV-2 outbreak.

---

**Supplemental figure 1.**
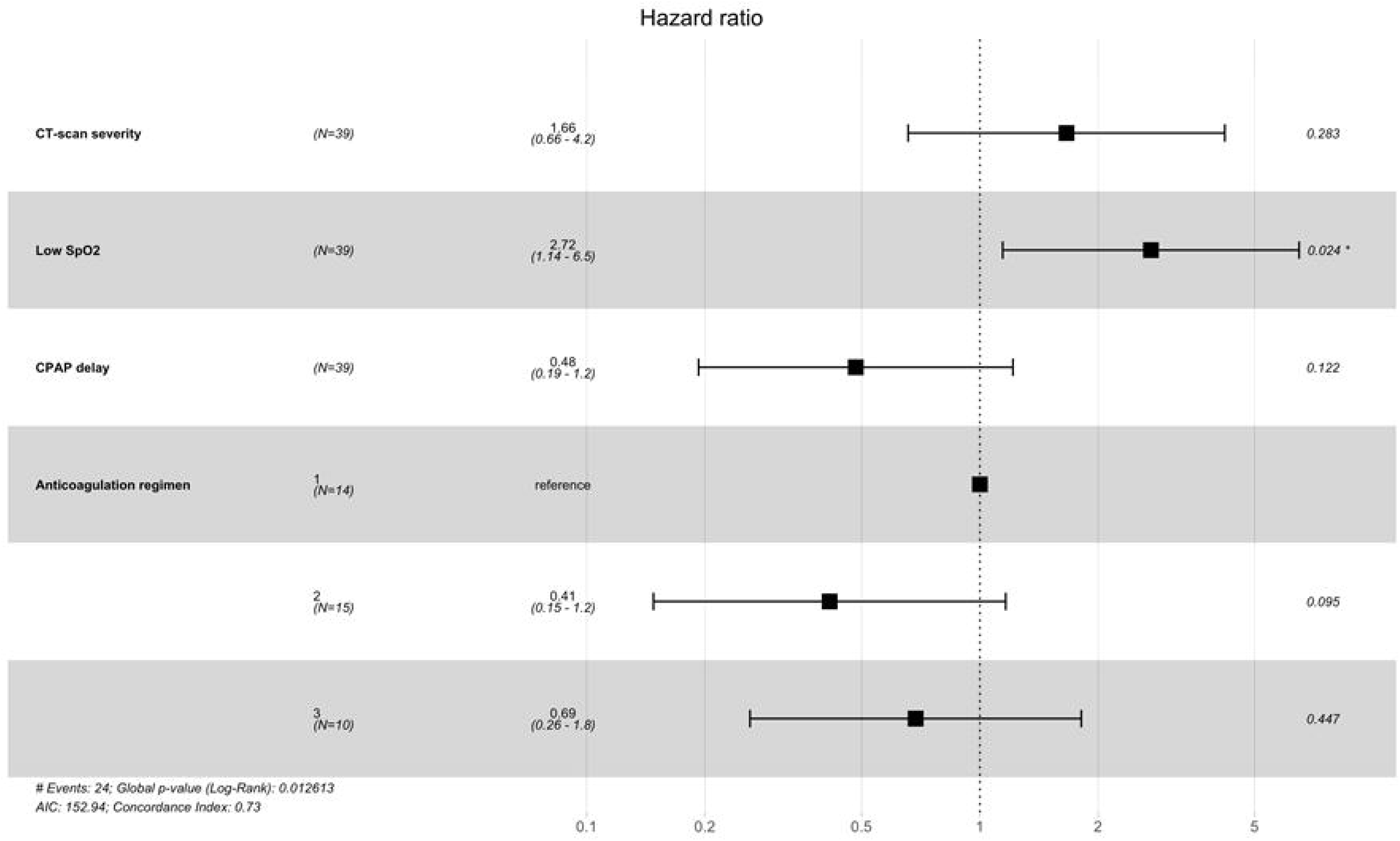
Hazard ratio of intubation adjusted for CT-scan severity (more or less of 50 % of lung involved by SARS—CoV2 induced lesions), low saturation (Sp02, < 92 % or > 92 %), delay in days between hospitalisation and CPAP initiation (two groups based on the median value of CPAP delay), use of anticoagulant treatment grouped by dosage: simple dose prophylaxis (1), double dose prophylaxis (2) or curative treatment (3). *P* values are indicated as the result of likelihood-ratio test. The validity of the proportional hazards assumption was tested using cox.zph() function in R (P values > 0.05) and by visualisation of Schoenfeld residuals.

